# Integrated clinical risk prediction of type 2 diabetes with a multifactorial polygenic risk score

**DOI:** 10.1101/2024.08.22.24312440

**Authors:** Scott C. Ritchie, Henry J. Taylor, Yujian Liang, Hasanga D. Manikpurage, Lisa Pennells, Carles Foguet, Gad Abraham, Joel T. Gibson, Xilin Jiang, Yang Liu, Yu Xu, Lois G. Kim, Anubha Mahajan, Mark I. McCarthy, Stephen Kaptoge, Samuel A Lambert, Angela Wood, Xueling Sim, Francis S. Collins, Joshua C. Denny, John Danesh, Adam S. Butterworth, Emanuele Di Angelantonio, Michael Inouye

## Abstract

Combining information from multiple GWASs for a disease and its risk factors has proven a powerful approach for development of polygenic risk scores (PRSs). This may be particularly useful for type 2 diabetes (T2D), a highly polygenic and heterogeneous disease where the additional predictive value of a PRS is unclear. Here, we use a meta-scoring approach to develop a metaPRS for T2D that incorporated genome-wide associations from both European and non-European genetic ancestries and T2D risk factors. We evaluated the performance of this metaPRS and benchmarked it against existing genome-wide PRS in 620,059 participants and 50,572 T2D cases amongst six diverse genetic ancestries from UK Biobank, INTERVAL, the All of Us Research Program, and the Singapore Multi-Ethnic Cohort. We show that our metaPRS was the most powerful PRS for predicting T2D in European population-based cohorts and had comparable performance to the top ancestry-specific PRS, highlighting its transferability. In UK Biobank, we show the metaPRS had stronger predictive power for 10-year risk than all individual risk factors apart from BMI and biomarkers of dysglycemia. The metaPRS modestly improved T2D risk stratification of QDiabetes risk scores for 10-year risk prediction, particularly when prioritising individuals for blood tests of dysglycemia. Overall, we present a highly predictive and transferrable PRS for T2D and demonstrate that the potential for PRS to incrementally improve T2D risk prediction when incorporated into UK guideline-recommended screening and risk prediction with a clinical risk score.

## Introduction

The global prevalence of type 2 diabetes (T2D) has quadrupled in the last 30 years, affecting approximately 508 million adults globally in 2021, with prevalence expected to increase a further 60% by 2050^1,2^. The risk of developing T2D is determined by a complex interplay of lifestyle, environmental, and genetic factors^3^. Genetic studies have estimated the heritability of T2D to be 69% among adults 35–60 years of age^4^ and genome-wide association studies (GWAS) have thus far identified 611 genomic loci associated with T2D risk^5^.

Polygenic risk scores (PRS) have emerged as a powerful tool for aggregating genomic associations into a single score quantifying an individual’s genetic predisposition to disease^6–8^. As they are based on the germline genome, which is stable throughout the life-course, a key advantage of PRS in comparison to other risk factors is early risk prediction. PRS can be used to predict disease risk at any point in a lifetime, including decades before lifestyle and environmental risk factors for T2D manifest, and it has been widely shown that risk prediction models can improve their ability to predict risk when PRS are integrated with commonly used risk predictors^6–8^. Numerous T2D PRS have been constructed to date, with 134 PRS from 40 studies published in the Polygenic Score (PGS) Catalog^9^ at the time of writing.

Most PRS have been developed using a single source of GWAS summary statistics. However, substantial improvements in prediction have been found by studies combining multiple sources of GWAS summary statistics during PRS development^10–13^. Improvements in PRS performance have been obtained both by combining information from multiple GWASs or PRSs from the disease of interest^10^ as well as by incorporating information from GWASs for disease risk factors^11–13^. Yet, PRS tailored specifically for T2D using this strategy are currently lacking. It is unclear to what extent this will improve predictive performance, transferability, and/or add value beyond existing clinical risk scores.

Here we utilize ancestrally diverse GWAS summary statistics from ten T2D GWAS and 34 T2D risk factor GWASs to develop a PRS for T2D. This new T2D metaPRS is externally validated and compared with previously published PRS in six diverse genetic ancestries from four large independent cohorts/biobanks: UK Biobank^14,15^, INTERVAL^16,17^, the All of Us research program^18–20^, and the Singapore Multi-Ethnic Cohort^21^. We further compare the T2D metaPRS and assess its added value to conventional risk factors and QDiabetes risk prediction scores^22^ for 10-year T2D risk prediction in UK Biobank.

## Results

### Study participants

A schematic of the overall study design is shown in **Figure 1**. After filtering, in total we analysed data from 620,059 participants, including 50,572 T2D cases, across the four study cohorts (**Methods**). Participants were grouped into genetic clusters using principal components analysis and assigned ancestry labels 1KG-EUR-like, 1KG-AFR-like, 1KG-AMR-like, 1KG-SAS-like, and 1KG-EAS-like based on their similarity to 1000 genomes (1KG) reference panel superpopulations^23^ following the 2023 National Academies guidelines on using population descriptors in genetics and genomics research^24^. Importantly, these labels seek to recognize (1) that genetic ancestries are distinct from and frequently do not overlap with ethnic and cultural identities, (2) these groupings are defined based on genetic similarity to arbitrary sets of labelled reference individuals, and (3) these groupings, while useful tools for statistical analyses, are artificial and do not represent the continuum of genetic diversity that exists in the human population^24^. Ethnic Malays in the Singapore Multi-Ethnic Cohort were handled separately as their genetic ancestries are not well represented by the 1KG reference panel, e.g. they do not cluster with either the 1KG EAS or SAS reference populations^25^. For consistency with the other genetic ancestry labels, here we assign the label “Austronesian-like” (ASN-like) to reflect their ancestral population histories^25^. Characteristics of each genetic ancestry and cohort are described in **Table S1**.

**Figure 1:**
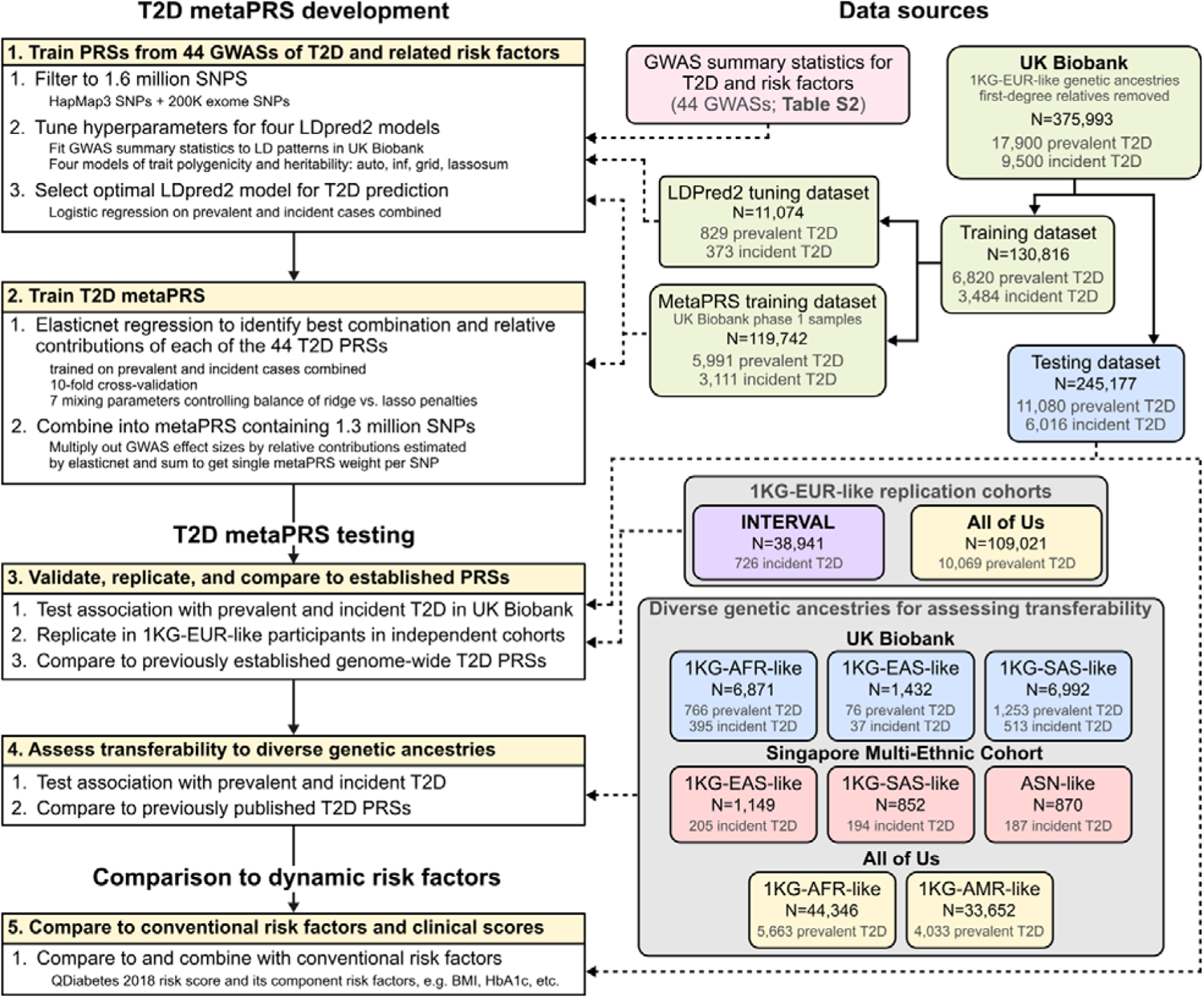
Study Design. Acronyms are as follows. T2D: type 2 diabetes. PRS: polygenic risk scores. SNPs: single nucleotide polymorphisms. LD: linkage disequilibrium. GWAS: genome-wide association study. 1KG-AFR-ike, 1KG-AMR-like, 1KG-EAS-like, 1KG-EUR-like, 1KG-SAS-like: genetic ancestry labels, defined based on clustering of participants by genetic principal components and the similarity of those clusters to 1000 Genomes reference panel superpopulations following the 2023 National Academies guidelines on using population descriptors in genetics and genomics research. ASN-like: genetic ancestry label chosen for ethnic Malays in the Singapore Multi-Ethnic cohort to represent their ancestral population history due to lack of representation of Austronesian populations in the 1000 Genomes reference panel.

### Derivation of a metaPRS for type 2 diabetes

To develop the metaPRS for type 2 diabetes we split unrelated 1KG-EUR-like UK Biobank participants into a PRS training dataset (N=130,816; 10,304 T2D cases) and a PRS testing dataset (N=245,117; 17,096 T2D cases) (**Methods**, **Figure 1**). To train the metaPRS, we used our previously described meta-scoring approach^11^, which leverages information from PRS trained on multiple GWAS of the target disease and its risk factors (**Methods**). Summary statistics were all obtained from contemporary GWASs that did not include UK Biobank participants (**Table S2**). We trained 44 PRSs to predict T2D using LDpred2^26^ and summary statistics from 10 GWAS of T2D across diverse ancestries and 34 GWAS for T2D risk factors (**Figure S1**, **Table S3**). The 44 PRSs were subsequently combined into a single metaPRS using elasticnet logistic regression^27^ with 10-fold cross validation in the training dataset (**Figure S2**, **Table S4**). The T2D metaPRS comprising 1.3 million SNPs is made available on the PGS Catalog^9^ with accession PGS004923.

### The metaPRS improves risk prediction of type 2 diabetes compared with other PRSs

Using the independent 1KG-EUR-like UK Biobank testing dataset of 245,117 participants, we next quantified the performance of the metaPRS for predicting prevalent T2D case status (11,080 cases) at baseline and for predicting risk of incident T2D (6,016 cases from hospital episode statistics) over 10-years of follow-up via survival analysis. All associations were adjusted for age, sex, and 20 genetic principal components (PCs). Prevalent and incident T2D cases in UK Biobank were analysed separately due to substantial differences in case identification^28^ (**Methods**). T2D is primarily diagnosed by primary care physicians, however less than half the participants had linked primary care records available. Prevalent cases were identified using a combination of self-reported diabetes diagnoses, prescription medication usage, and retrospective hospital records, whereas identification of incident T2D cases relied solely on hospital records. The metaPRS was associated with prevalent T2D with an odds ratio of 2.30 (95% CI: 2.26–2.35) per standard deviation of the metaPRS, with an area under the receiver-operating characteristic curve (AUC) of 0.777 (95% CI: 0.772–0.781). The metaPRS was associated with incident T2D with a hazard ratio (HR) of 1.80 (95% CI: 1.75–1.85) per standard deviation of the metaPRS, with a C-index of 0.719 (95% CI: 0.713–0.725). When compared to other PRS (**Table S6**) that could be evaluated in 1KG-EUR-like UK Biobank samples (i.e., did not include UK Biobank GWAS in PRS training), the metaPRS had the strongest associations with both prevalent and incident T2D (**Figure 2 A–B, Table S5**).

**Figure 2:**
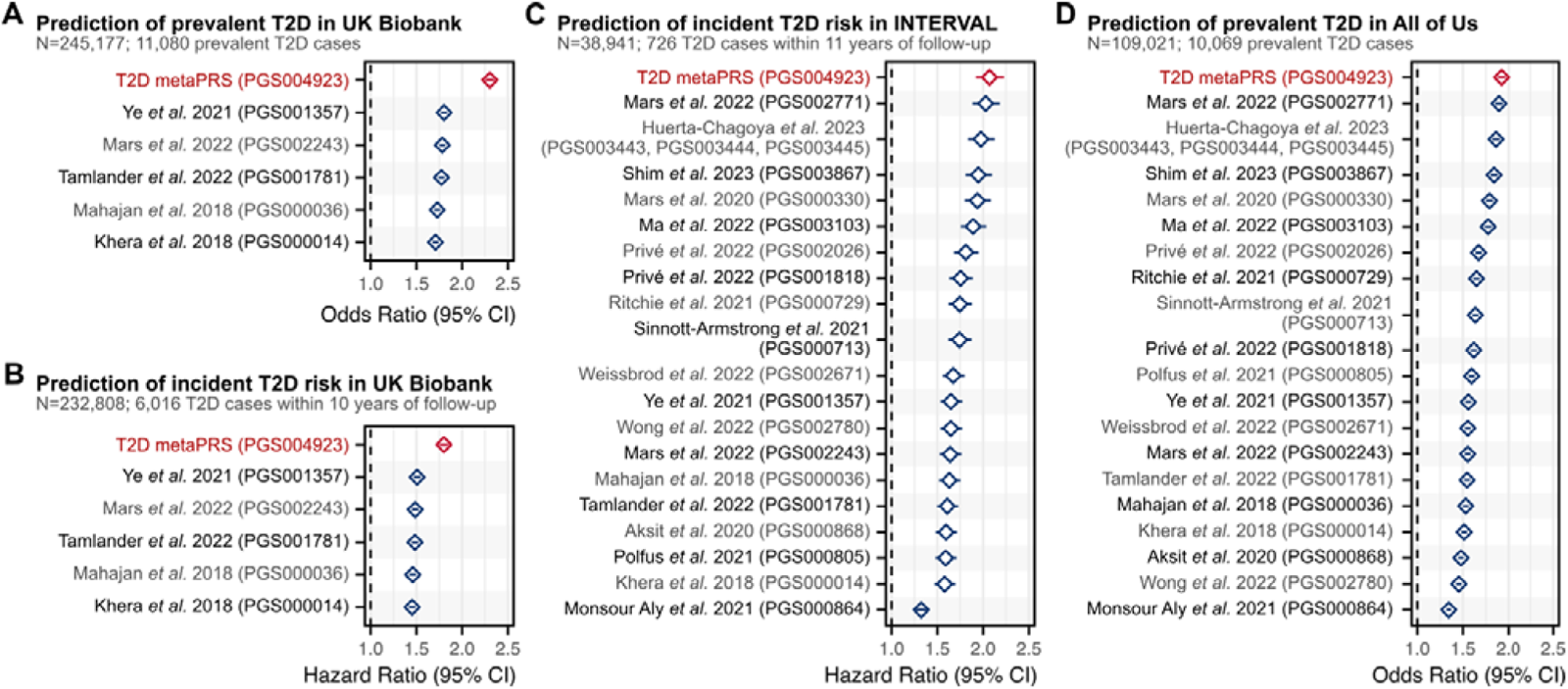
Comparison of T2D PRSs in people of 1KG-EUR-like genetic ancestries across three cohorts. Comparison of PRSs for association with prevalent T2D status or time-to-onset of incident T2D in participants of 1KG-EUR-like genetic ancestries from the UK Biobank, INTERVAL, and All of Us research program cohorts. In UK Biobank incident and prevalent T2D were analysed separately due to significant difference in phenotype severity (**Methods**). Analyses of UK Biobank excluded participants used for metaPRS training, and PRSs derived from GWAS performed in UK Biobank samples. The limited number of PRSs tested in UK Biobank compared to INTERVAL and All of Us reflects that the majority of contemporary PRSs utilize GWAS performed in UK Biobank samples for PRS development. PRSs were adjusted for 20 genetic principal components in each cohort prior to model fitting. Diamonds show the odds ratios or hazard ratios, and horizontal bars show the 95% confidence intervals. Odds ratios and hazard ratios are per standard deviation increase in the respective PC-adjusted PRS. Logistic and Cox proportional hazards regressions were adjusted for age, sex, and cohort specific covariates (e.g., assessment centre). Odds ratios and hazard ratios are detailed in **Table S6.** Details on comparison PRSs are provided in **Table S5**.

To replicate the metaPRS and compare to contemporary PRS trained using 1KG-EUR-like UK Biobank GWAS, we analysed data from a combined 1KG-EUR-like 147,962 participants (10,795 T2D cases) from the INTERVAL cohort^16,17^ and the All of Us research program^18–20^ (**Figure 2C–D**, **Table S5**). In INTERVAL, the metaPRS was associated with incident T2D with a HR of 2.07 (95% CI: 1.92–2.23) and a C-index of 0.774 (95% CI: 0.758–0.790). In All of Us, the metaPRS was associated with prevalent T2D with an odds ratio of 1.92 (95% CI: 1.88–1.97) and an AUC of 0.737 (95% CI: 0.732–0.742). Importantly, when compared to other genome-wide PRSs (**Table S6**), the metaPRS was the strongest predictor of T2D in both cohorts. In both cohorts the second strongest PRS was that of Mars *et al.* 2022 (PGS002771)^29^, which had a HR of 2.03 (95% CI: 1.88–2.18) and C-index of 0.772 (95% CI: 0.756–0.788) in INTERVAL and an odds ratio of 1.89 (95% CI: 1.85–1.94) and AUC of 0.735 (95% CI: 0.730–0.740) in All of Us. Furthermore, the relative performance of PRSs was remarkably consistent across both INTERVAL and All of Us (**Figure 2C–D**).

### Transferability of the metaPRS across diverse genetic ancestries

To assess the transferability of the metaPRS and other T2D PRS beyond 1KG-EUR-like genetic ancestries, we analysed data from a combined 96,164 participants (12,377 T2D cases) clustering into five genetic ancestries (1KG-AFR-like, 1KG-AMR-like, 1KG-SAS-like, 1KG-EAS-like, and ASN-like) from the UK Biobank^16,17^, the All of Us research program^18–20^, and the Singapore Multi-Ethnic Cohort^21^. As expected, we observed considerable heterogeneity in both absolute and relative strength of associations of PRS across genetic ancestries and cohorts (**Table S7**). Notably, no single PRS emerged as the most predictive, even within any given genetic ancestry group: the top PRS was both ancestry and cohort specific (**Figure 3**). When comparing relative effect sizes across cohorts and genetic ancestries, four PRS emerged as the most consistent top performers: our metaPRS, along with PRSs from Huerta-Chagoya *et al.* 2023 (weighted sum of PGS003443, PGS003444, and PGS003445; **Methods**)^30^, Shim *et al.* 2023 (PGS003867)^31^, and Mars *et al.* 2022 (PGS002771)^29^ (**Figure 4**). As expected^32,33^, the predictive power of all tested PRSs weakened as genetic ancestries diverged from 1KG-EUR-like: from a maximum odds ratio of 2.30 (95% CI: 2.26–2.35) for any PRS in 1KG-EUR-like samples (**Table S6**), to 1.91 (95% CI: 1.78–2.05) in 1KG-SAS-like, 1.90 (95% CI: 1.48–2.45) in 1KG-EAS-like, 1.77 (95% CI: 1.71–1.84) in 1KG-AMR-like, 1.71 (95% CI: 1.43–2.06) in ASN-like, and 1.37 (95% CI: 1.33–1.41) in 1KG-AFR-like (**Table S7**).

**Figure 3:**
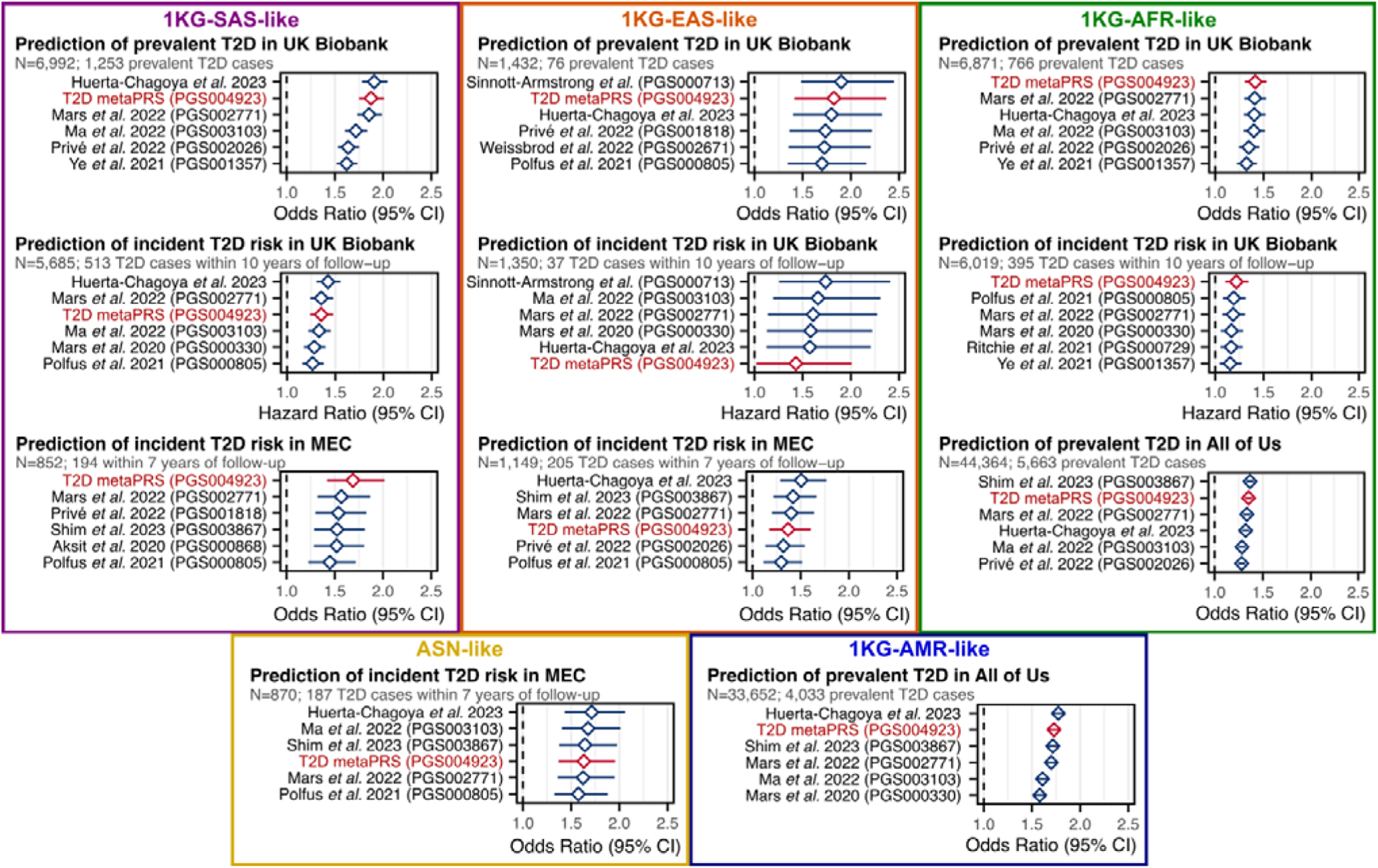
Top six T2D PRSs in people of five diverse ancestry groups across three cohorts. Comparison of PRSs for association with T2D status or time-to-onset in participants clustering into five diverse ancestry groups from UK Biobank, the All of Us research program cohort, and the Singapore Multi-Ethnic Cohort (MEC). Participants in each cohort were clustered by genetic similarity with the 1KG reference population participants (**Methods**), except for ethnic Malays in the MEC study, as their genetic ancestries are distinct from and not represented by 1KG reference populations, and here labelled as Austronesian (ASN)-like to reflect their ancestral population histories. The top six PRS for each cohort and genetic ancestry group are shown; in UK Biobank incident and prevalent T2D were analysed separately due to significant difference in phenotype severity (**Methods**). Odds ratios and hazard ratios for all tested PRS are detailed in **Table S7**. PRSs were adjusted for 20 genetic principal components in each cohort prior to model fitting. Diamonds show the odds ratios or hazard ratios, and horizontal bars show the 95% confidence intervals. Odds ratios and hazard ratios are per standard deviation increase in the respective PC-adjusted PRS. Logistic and Cox proportional hazards regressions were adjusted for age, sex, and cohort specific covariates (e.g., assessment centre). Logistic regression was used to assess associations with incident T2D in MEC as time to T2D onset (or T2D-free survival) was not available due to the heterogeneity of incident T2D ascertainment (**Methods**). Note the Shim *et al.* 2023 PRS (PGS003867) could not be tested in UK Biobank as it was derived from multi-ancestry GWAS performed in UK Biobank samples.

**Figure 4:**
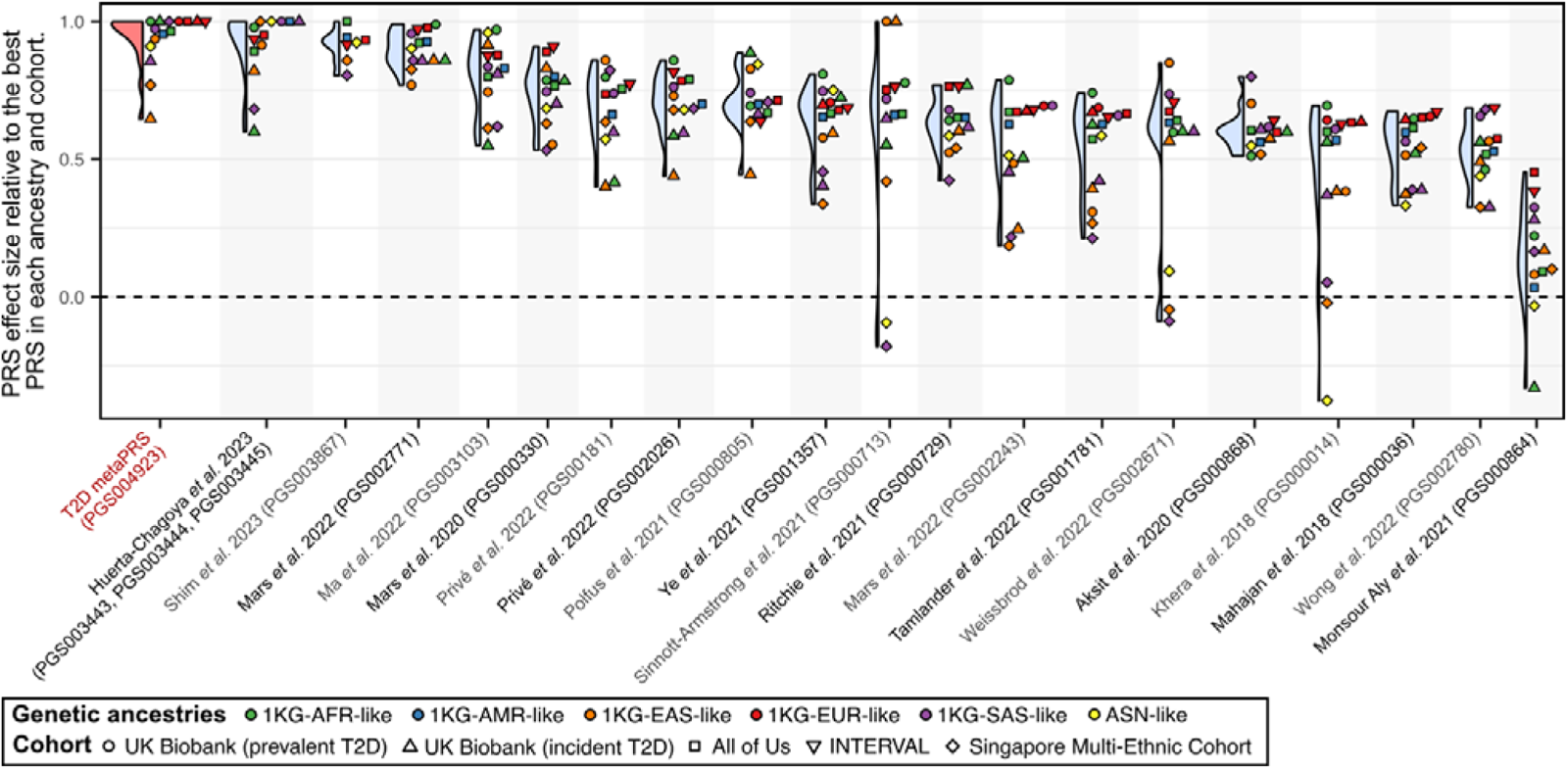
Relative rank of PRSs by effect size within each genetic ancestry and cohort. Violin and dotplot comparing PRS effect size relative to the PRS with the maximum effect size within each cohort and genetic ancestry group. A score of 1.0 is given to the PRS with the maximum odds ratio or hazard ratio in each cohort and genetic ancestry combination. Other PRSs were then assigned a relative prediction value based on the ratio of their log odds ratio (or log hazard ratio) to that of the strongest PRS. PRS are ordered left to right based on their median score.

### Comparison to conventional risk factors and QDiabetes risk scores

We compared the metaPRS to established T2D risk factors and QDiabetes^22^, a 10-year T2D risk prediction score recommended to clinicians by the UK’s National Institute for Health and Care Excellence (NICE) guidelines for T2D prevention^34^ and National Health Service (NHS) health check best practice guidance^35^. For this, we utilize a subset of 190,293 1KG-EUR-like UK Biobank participants (4,064 incident T2D cases) with risk factor information required for QDiabetes risk score calculation (**Figure 5A**, **Table S8**). The C-index for the metaPRS (C-index: 0.716; 95% CI: 0.708–0.723) was larger than for all individual risk factors—including family history (C-index: 0.687; 95% CI: 0.679–0.695)— except for body mass index (BMI) (C-index: 0.780; 95% CI: 0.773–0.787) and glycated haemoglobin (HbA1c) (C-index: 0.826; 95% CI: 0.819–0.833).

**Figure 5:**
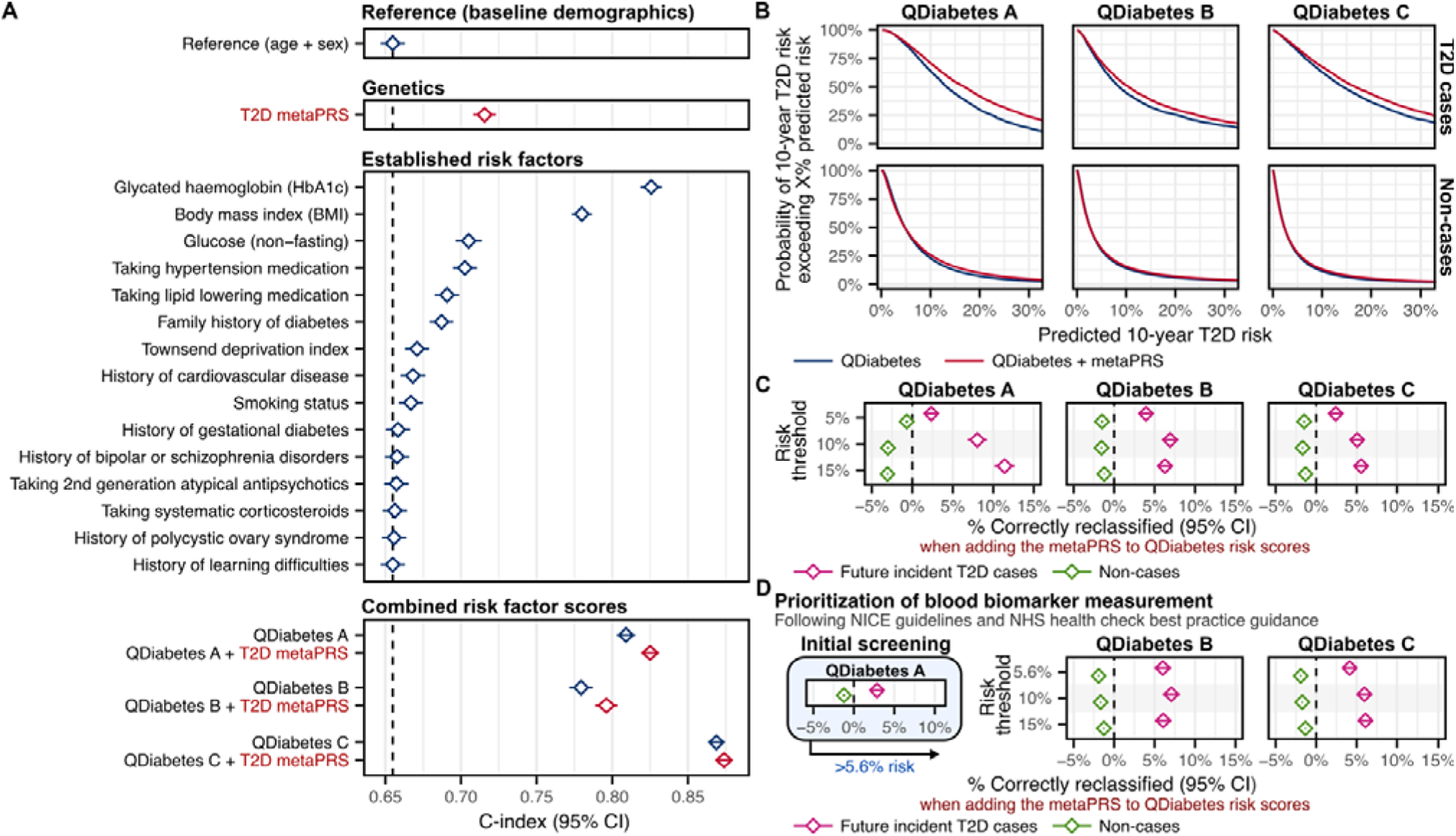
Comparison to established risk factors and risk prediction scores. **A)** Comparison of C-index over age and sex alone for the metaPRS to individual type 2 diabetes risk factors and 10-year type 2 diabetes risk prediction scores (QDiabetes) in 190,293 1KG-EUR-like UK Biobank participants (4,064 incident T2D cases). The QDiabetes 2018 model A score is calculated from all listed individual risk factors, excepting glucose and HbA1c. The QDiabetes 2018 model B and model C scores additionally incorporates fasting glucose and HbA1c respectively. Note UK Biobank participants are non-fasting leading to likely underestimation of QDiabetes B. For comparison purposes the set of participants analysed here was selected as the subset in which the QDiabetes 2018 model C risk score could be computed (complete risk factor information, with height between 1.4 and 2.1 meters, weight ≤ 180 kg, and HbA1c between 15 and 48 mmol/mol). Diamonds show the C-index and horizontal bars show the 95% confidence intervals. C-indices and individual risk factor hazard ratios are detailed in **Table S8**. **B)** Probability of predicted 10-year risk exceeding X% when using QDiabetes risk with or without the T2D metaPRS. Probabilities were calculated as one minus the empirical cumulative distributive function across cases and non-cases combined. Probability curves extend to the right of each plot up, to 100% predicted risk, but are truncated here for clarity. **C)** Categorical net reclassification improvement (NRI) when adding the metaPRS to QDiabetes risk scores for stratifying participants into high and low risk groups at varying risk thresholds. % correctly reclassified: net % of cases that were correctly reclassified from the low-risk group into the high-risk group when adding the metaPRS (pink) or the net % of non-cases that were correctly reclassified from the high-risk group into the low-risk group when adding the metaPRS (green). 95% confidence intervals were estimated via a bootstrap sampling procedure with 1000 bootstraps. Diamonds show the net % correctly reclassified and horizontal bars show the 95% confidence intervals. Categorical NRI details and numbers allocated to each risk category are provided in **Table S9**. **D)** Categorical NRI when incorporating the metaPRS into a two-stage procedure in which QDiabetes model A is used to prioritize potential high-risk individuals for fasting glucose or HbA1c blood tests for subsequent risk prediction and stratification. Categorical NRI details and numbers allocated to each risk category are provided in **Table S10**.

When added to QDiabetes and its model variants (A, B and C), the metaPRS significantly improved 10-year T2D risk prediction (**Figure 5A**, **Table S8**). The basic QDiabetes score (model A), which incorporates all risk factors that do not require taking a blood sample (**Methods**), had a C-index of 0.808 (95% CI: 0.802–0.814). Adding the metaPRS to QDiabetes model A increased the C-index by 0.016 (95% CI: 0.013– 0.019; P-value: 6×10^−34^) yielding a total C-index of 0.824 (95% CI: 0.818–0.830). The QDiabetes score incorporating fasting glucose (model B) had a C-index of 0.773 (95% CI: 0.765–0.781). The substantially lower C-index for model B compared to model A of the QDiabetes score can be explained by the non-fasting status of UK Biobank participants, which would lead to overestimation of risk for those who have recently eaten (i.e. have higher glucose). Adding the metaPRS to QDiabetes model B led to a similar increase in C-index compared to model A, with a ΔC-index of 0.019 (95% CI: 0.015–0.022; P-value: 6×10^−27^), yielding a total C-index of 0.790 (95% CI: 0.783–0.798). The QDiabetes score incorporating HbA1c (model C) had the largest C-index of 0.866 (95% CI: 0.861–0.872). Addition of the metaPRS to this model led to a smaller, but still statistically significant, increase in C-index (ΔC-index: 0.005; 95% CI: 0.004–0.006; P-value: 4×10^−15^), yielding a total C-index of 0.871 (95% CI: 0.866–0.877).

When incorporating the metaPRS into absolute risk predictions made by QDiabetes risk scores (**Figure 5B, Supplementary Methods**) we observed significant improvements in risk stratification at varying risk thresholds (5%, 10%, 15%) for all QDiabetes model variants (**Figure 5C**, **Table S9**). Consistent with the above, improvements in risk stratification were strongest when adding the metaPRS to QDiabetes score model A. Using a threshold of 10% absolute risk, we observed a net 8.02% improvement (95% CI: 6.83%– 9.22%; P-value: 1×10^−39^) in classification of future incident T2D cases as high risk when adding the metaPRS to QDiabetes score model A. Among the 4,064 incident T2D cases, the number of cases correctly identified as high risk increased from 2,509 to 2,853 (an additional 11.52% of cases correctly identified as high risk) with 142 cases (3.50%) incorrectly reclassified as low risk (net improvement of 8.02%). Net improvements in risk stratification of cases using a 10% risk threshold were 6.92% (95% CI: 5.88%– 7.96%; P-value: 6×10^−39^) for QDiabetes model B and 5.07% (95% CI: 4.13%–6.02%; P-value: 8×10^−26^) for QDiabetes model C respectively. Modest, but statistically significant, increases in the number of non-cases incorrectly classified as high risk were also observed at all tested risk thresholds (**Figure 5C**, **Table S9**). With the 10% risk threshold, the net number of non-cases incorrectly classified as high-risk increased by 3.01% (95% CI: 2.87%–3.14%; P-value < 1×10^−300^) when adding the metaPRS to QDiabetes model A, by 1.56% (95% CI: 1.46%–1.66%; P-value: 3×10^−217^) when adding the metaPRS to QDiabetes model B, and by 1.67% (95% CI: 1.59%–1.76%; P-value < 1×10^−300^) when adding the metaPRS to QDiabetes model C.

### Improvements in risk stratification and screening following UK guidelines

NICE guidelines for T2D prevention^34^ and NHS health check best practice guidance^35^ recommend using the basic QDiabetes score (model A) to prioritize potential high risk individuals (>5.6% risk) for fasting glucose or HbA1c blood tests, which can then be used subsequently to enhance risk prediction via QDiabetes models B and C^22^. When modifying the initial screening step by adding the metaPRS to QDiabetes model A, the number of participants with >5.6% risk prioritized for blood tests increased from 75,153 (3,396 incident T2D cases) to 77,495 (3,517 incident T2D cases); yielding a similar number to follow-up with blood tests per T2D event (number needed to screen; NNS) of 22.13 vs. 22.03 respectively (ΔNNS: −0.10, 95% CI: −0.33–0.14, P-value: 0.14). Net improvements in risk stratification of T2D cases after applying QDiabetes model B or C to these prioritized individuals (**Figure 5D**) of 4%–7% were observed (**Figure 5D**, **Table S10**), similar to those observed above when systematically assessing all participants with QDiabetes models B or C. Likewise, a modest but statistically significant increase in the net number of non-cases incorrectly classified as high risk of 1%–2% was also observed when incorporating metaPRS into both stages of the guideline-recommended screening procedure (**Figure 5D**, **Table S10)**.

When applying the QDiabetes author-recommended^22^ risk-threshold of 14.3% on QDiabetes model C after using QDiabetes model A to prioritize individuals for HbA1c measurement (**Table S10**), a total of 10,745 participants (5.6%) were classified as high-risk, including 1,906 of the future T2D cases (46.9%). When adding the metaPRS to both the initial screening with QDiabetes model A and subsequent risk prediction with QDiabetes model C, these increased to a total of 13,564 participants (7.1%) and 2,167 cases (53.3%) classified as high-risk, yielding a net absolute gain in case classification of 6.41% (95% CI: 5.43%–7.38%; P-value: 7×10^−38^) and a net increase in the number of non-cases incorrectly classified as high-risk of 1.37% (95% CI: 1.30%–1.45%; P-value: 2×10^−279^). When considering the ratio of total interventions recommended per T2D case among those at high-risk (number needed to treat; NNT), a modest but statistically significant increase in NNT from 5.64 to 6.26 (ΔNNT: 0.62, 95% CI: 0.50–0.74, P-value: 2×10^−24^) was observed.

## Discussion

In this study, we developed a PRS for T2D based on summary statistics from 44 GWASs for T2D and its risk factors. We quantified the predictive power of the T2D metaPRS by performing the broadest benchmarking of genome-wide T2D PRS to date (i.e. over half a million participants from six diverse genetic ancestry groups from four population-based cohorts from the UK, US, and Singapore). In benchmarking, we demonstrated that the T2D metaPRS is the most predictive PRS for T2D in European genetic ancestries and had comparable performance to the top ancestry- and cohort-specific PRS, highlighting its transferability. We further compared the T2D metaPRS to established non-genetic risk factors and quantified its added value in combination with 10-year risk prediction scores in the context of current UK guidelines^34,35^.

Transferability is a major challenge for PRS development and a barrier to PRS utility and equitable clinical application. Data availability has meant PRS have predominantly been developed using GWAS from European genetic ancestries^36,37^. This risks exacerbating health disparities as PRSs have shown reduced predictive performance in individuals of non-European and complex genetic ancestries^32,33^, whom make up the majority of the global population. In our systematic benchmarking, the majority of PRSs showed reduced performance relative to other PRSs when tested outside of the genetic ancestries used in their development, with worsening performance as the continuum of genetic ancestries diverged. PRSs developed with ancestry-specific data were also frequently out-performed by out-of-ancestry or multi-ancestry PRSs, likely due to differences in available sample sizes. Surprisingly, we also found that within non-1KG-EUR-like ancestries there was no single maximally predictive PRS in each ancestry group; top PRSs were both ancestry- and cohort-specific. Moreover, the absolute magnitude of odds ratios weakened as genetic ancestries diverged from 1KG-EUR-like, including for PRSs developed in non-1KG-EUR-like samples using non-1KG-EUR-like GWAS summary statistics. Our results add to the body of evidence highlighting the need for recruitment of participants from globally and genetically diverse ancestries as part of large biobanks and cohorts, including and beyond high-income countries^36,37^. Our results further highlight that the relative performance of PRSs can also differ considerably between cohorts even within the same genetic ancestry group, suggesting heterogeneity in environment or phenotype definition can also impact transferability^38^.

When compared to established risk factors, the metaPRS had stronger predictive power for 10-year risk than all conventional risk factors, apart from BMI and biomarkers of dysglycemia, and captured residual risk not quantified by these risk factors. The metaPRS also provided a modest, but statistically significant, improvement over the QDiabetes risk scores combining established risk factors for both risk discrimination and risk stratification at varying risk thresholds. Improvements from the metaPRS were less than those from measurement and addition of blood biomarkers of dysglycemia (e.g. HbA1c), consistent with a previous study of T2D PRSs in a population cohort of British Pakistanis and Bangladeshis^39^.

However, blood tests for dysglycemia are not routinely conducted in asymptomatic individuals; UK guidelines recommend using readily available lifestyle and medical history information to identify high-risk individuals (i.e. using QDiabetes model A) for follow-up testing of fasting glucose or HbA1c blood tests for T2D diagnosis^34,35^. QDiabetes models B and C have been developed with a view to enhancing risk prediction in those found to not be diabetic after follow-up blood tests^22^. PRSs may one day be included among readily available factors for risk screening as they require a one-off blood sample for genotyping which may be obtained at any time during a person’s life, for example via initiatives like the UK Newborn Genome Screening Programme^40^.

Here, we show that, if genotypes are already available, the metaPRS can enhance this initial screening step: increasing from 84% to 87% the number of future diabetics revealed to be at elevated risk by blood testing, with a similar NNS of ∼22. The metaPRS also improved subsequent risk prediction, increasing the number of T2D cases classified as high-risk by 6.4% when used alongside HbA1c with QDiabetes model C, with a modest increase in the NNT by 0.62 from 5.64 to 6.26.

Our study has limitations. Firstly, while we utilized multi-ancestry GWAS summary statistics whenever available, a 1KG-EUR-like cohort was used for model training. As large and diverse training sets with T2D outcomes become available, future studies can utilize highly diverse cohorts for PRS training alongside genetically diverse GWAS, thus resulting in more powerful, and more portable, T2D PRS. The UK Biobank samples used for analysis of established risk factors and QDiabetes risk scores differs from the wider UK population in several key respects. UK Biobank participants are healthier than the general UK population^14^ and thus prevalence will be higher for dichtomous risk factors (e.g. medical history) and distributions will be narrower and/or shifted for continuous risk factors (e.g. BMI). Participants were also non-fasting, confounding risk predictions made by QDiabetes model B which relies on fasting glucose^22^. We also expect risk stratification to substantially differ from the general population, as T2D is primarily diagnosed by primary care physicians, whereas incident T2D case identification in UK Biobank relied on hospital records since less than half the cohort has linked primary care records available. Likewise, the high-risk sub-population assessed subsequent to screening did not exclude those with undiagnosed T2D as diagnosis was not possible as this required fasting glucose or repeated HbA1c measures^34,35^. Our analyses were also restricted to genetically homogenous 1KG-EUR-like participants self-reporting as White British due to the much smaller sample sizes available for other ethnic and ancestry groups and expected confounding from population stratification that would be introduced if assessing the metaPRS in the pooled multi-ethnic and multi-ancestry sample^41,42^. Despite these limitations, our analyses nevertheless indicate that incorporating our T2D metaPRS could modestly improve screening and risk stratification. Further studies in cohorts representative of the general UK population eligible for screening would be needed to accurately quantify the precise added benefits of PRS to screening and 10-year risk prediction.

Overall, our study presents a new T2D PRS that is highly predictive across diverse genetic ancestries and cohorts, improves risk prediction when added to established risk factors in clinical risk scores for 10-year risk prediction of T2D, and has the potential to improve screening practices in the UK.

## Methods

### UK Biobank cohort

UK Biobank is a cohort of approximately 500,000 individuals with deep phenotyping, imputed genotypes, and electronic health record linkage^14,15^. Participants were members of the general UK population between 40 to 69 years of age identified and recruited through primary care lists and who accepted an invitation to attend one of 22 assessment centres across the UK between 2006 and 2010. Ethics were approved by the North West Multi-centre Research Ethics Committee (MREC) in the UK, and this study was undertaken under UK Biobank project #7439. Participants gave informed and broad consent for health-related research.

Recruitment included standardized questionnaires on socio-demographics, ethnicity, lifestyle factors, and personal- and family-medical history. Physical measurements including height, weight, body fat percentage, and systolic blood pressure were also taken at assessment, and blood samples taken for genotyping and quantification of molecular phenotypes. Participants were also linked to national death and cancer registries as well as hospital episode statistics. Participants were genotyped on UK BiLEVE arrays and UK Biobank Axiom arrays and imputed to the 1000 genomes, UK10K, and Haplotype Reference Consortium panels^43^ using human genome build GRCh37^15^. Participants were filtered to a set of unrelated individuals (kinship < 0.0884) identified using kinship estimates^44^ supplied by UK Biobank^15^.

For the primary analyses of metaPRS derivation and validation we restricted analyses to the “White British” cohort defined by UK Biobank based on self-reported ethnicity (data-field #21000) combined with genetic principal components^15^. For consistency with other study cohorts and following the 2023 National Academies guidelines on using population descriptors in genetics and genomics research^24^ we assigned this group the genetic ancestries label 1KG-EUR-like. For analyses assessing PRS transferability we similarly defined genetically homogeneous populations using a combination of self-reported ethnicity and projection of genetic principle components to 1KG reference ancestral superpopulations^23^ using the KING software^44^. Participants were grouped into 1KG-SAS-like if they self-reported ethnicity as Indian, Pakistani, or Bangladeshi and their KING inferred ancestry was SAS with >95% probability. Participants were grouped into 1KG-EAS-like if they self-reported ethnicity as Chinese and their KING inferred ancestry was EAS with >95% probability. Participants were grouped into 1KG-AFR-like if they self-reported ethnicity as African, Caribbean, Black or Black British, or any other Black background and their KING inferred ancestry was AFR with >95% probability.

As linked primary care records are only available for less than half of UK Biobank participants, prevalent T2D status at baseline was adjudicated from a combination of retrospective hospital episode records, self-reported history of diabetes, and baseline medication using the Eastwood *et al.* algorithms^28^. Incident T2D cases were also ascertained following the Eastwood *et al.* algorithms^28^, on the basis of ICD-10 diagnosis coding E11 in either the hospital inpatient or death registry data. Onset of incident T2D was determined as the midpoint between the first hospital or death record with an ICD-10 E11 coding and the previous T2D-free record (hospital record without ICD-10 E11 coding or baseline assessment)^28^. Follow-up for incident T2D events was truncated on 1^st^ February 2020 to preclude potential confounding from SARS-CoV2 infection, exposure, or behavioural or environmental changes from pandemic lockdowns on metaPRS training.

### INTERVAL cohort

INTERVAL is a cohort of approximately 50,000 participants nested within a randomized trial studying the safety of varying frequency of blood donation^16,17^. Participants were blood donors aged 18 years and older (median 44 years of age; 49% women) recruited between June 2012 and June 2014 from 25 centres across England. Blood samples were taken at assessment and participants consented for broad health-related research^16,17^. Electronic health record linkage was available for a maximum of 11.1 years of follow-up (median 10.4 years). In total there were 38,949 participants who were diabetes free at baseline assessment with linked imputed genotypes and electronic health records. Ethics were approved for this study by the National Research Ethics Service (11/EE/0538).

Participants were genotyped using the Affymetrix UK Biobank Axiom arrays and imputed to the UK10K and 1000 Genomes panel using human genome build GRCh37. Notably, a key step in the genotype QC was exclusion of samples of non-European ancestry on the basis of genotype PCs^45^. For consistency with the other study cohorts and following the 2023 National Academies guidelines on using population descriptors in genetics and genomics research^24^ we assigned these participants the ancestry label 1KG-EUR-like.

Linked electronic health records from national hospital episode statistics were summarized into 301 endpoints from ICD-10 diagnosis codes using CALIBER rule-based phenotyping algorithms^46^ (https://www.caliberresearch.org/portal) prior to being made available to analysts. The closest matching CALIBER phenotype for T2D was for any diabetes; defined using ICD-10 codes E10–E14, G59.0, G63.2, H28.0, H36.0, M14.2, N08.3, or O24.0–O24.3. Participants with any diabetes history were excluded from the analysis. Incident diabetes events were treated as incident T2D for the purposes of analyses, consistent with the rarity of adult-onset type 1 diabetes. Onset of incident T2D was determined as the midpoint between the first diabetes event and the previous diabetes-free record (hospital record without a diabetes coding or baseline assessment).

### All of Us research program cohort

All of Us is a longitudinal cohort aiming to recruit one million participants from across the USA^18^. In the v7 data freeze, there were approximately 206,000 participants with deep phenotyping, whole genome sequencing, and electronic health record linkage^19^. Participants were members of the general USA population ≥18 years of age with recruitment focused on groups underrepresented in biomedical research^20^. Research was conducted on the All of Us Researcher Workbench under the guidelines defined by the All of Us Ethical Conduct of Research Policy.

Details of whole genome sequencing and quality control are described extensively in the All of Us Genomic Research Data Quality Report C2022Q4R9 at https://support.researchallofus.org/hc/en-us/articles/4617899955092-All-of-Us-Genomic-Quality-Report. Computation of kinship relatedness and clustering of participants by genetic similarity to 1KG AFR, EUR, and AMR reference ancestral superpopulations^23^ are also described in the report. Additional downstream quality control and filtering of sequence data is as described in Suzuki *et al.* 2024^5^. Briefly, related individuals were pruned to obtain a maximal independent set (kinship score > 0.1), and variants were filtered to high-quality SNPs with MAF > 1% or MAC > 100 in at least one of the genetic ancestry clusters. SNPs with MAF < 1% that deviated from Hardy-Weinberg equilibrium (P < 1×10^−6^) were removed. Principal components used for correction of population structure were calculated in each ancestry group separately using SNPs present in the 1000 Genomes project phase 3 release. Samples whose sex could not be imputed from genotypes were excluded.

Phenotyping of T2D case and control status was performed using the PheKB algorithm (https://phekb.org/phenotype/type-2-diabetes-mellitus) as described in Suzuki *et al.* 2024^5^. T2D cases were ascertained based on a combination of hospital diagnosis codes, prescription medication, and lab results from blood tests occurring prior to baseline sample assessment. Participants were considered controls if they had no history of any diabetes diagnoses, T2D medication, or abnormal glucose or HbA1c lab results. Participants with T1D or uncertain diabetes status were excluded from analysis.

### Singapore Multi-Ethnic Cohort

The Singapore Multi-Ethnic Cohort is a population-based cohort studying how genes and lifestyle influence disease risk differently in participants from three major ethnic groups in Singapore: Chinese, Indian, and Malay (https://blog.nus.edu.sg/sphs/population-studies/multi-ethnic-cohort-phase-1-mec1/)^21^. Participants were recruited between 2004 and 2010 and invited for follow-up assessment between 2011–2016 (mean follow-up 6.3 years). In total there were 2,871 participants with whole-genome sequencing who were disease-free at baseline. Written consent was obtained from all participants, and this study was approved by the National University of Singapore Institutional Review Board (reference codes: B-16-158 and N-18-059).

Details of whole genome sequencing and quality control are as previously described by the Singapore National Precision Medicine program strategy report^47^. Briefly, sequencing was performed to an average depth of 15x coverage. Reads were aligned with BWA-MEM v.0.7.17 and genotyped using GATK v.4.0.6.0. Variants were filtered to retain VQSR-PASS and non-STAR allele variants. Samples with call rate <95%, BAM cross-contamination rate >2%, BAM error rate > 1.5% were excluded. Genotypes with depth coverage (DP) < 5, genotype quality (GQ) < 20, or allele balance (AB) > 0.8 were set to null, and samples with abnormal ploidy excluded. Genetic variants were filtered to exclude those with robust, unified test for Hardy-Weinberg equilibrium (RUTH) P-value <0.01, a variant call rate <90%, being monomorphic, or having a minor allele count (MAC) <2 prior to phasing with Eagle version 2.4^43,48^. After quality control, the dataset included 39,967,216 genetic variants in 2,871 samples. Samples were clustered into three groups by genetic similarity using the k-means algorithm on the first 15 genetic principal components calculated on the verifyBamID2 variant panel (1KG phase 3)^49^.

Genetic ancestry labels for each cluster were based on the majority reported ethnicity in each group, here labelled as 1KG-EAS-like, 1KG-SAS-like, and ASN-like for consistency with other the study cohorts and following the 2023 National Academies guidelines on using population descriptors in genetics and genomics research^24^. The ASN-like label was used here to label the genetic cluster with Malay as the majority reported ethnicity, as their genetic ancestries were not well represented by either the EAS or SAS super populations in the 1KG reference panel^25^. The label Austronesian-like (ASN-like) was chosen to reflect the ancestral population histories of this group^25^.

Incident T2D (N=577) was ascertained as previously described^50^ through a combination of linkage to national healthcare records, self-reported medical history at follow-up assessment (either diagnosis from a primary care physician or current diabetes medication usage), or with blood biomarker concentrations indicative of diabetes following the American Diabetes Association criteria (fasting glucose ≥ 7 mmol/L or HbA1c ≥ 6.5% or random blood glucose ≥11 mmol/L)^51^.

### MetaPRS training

The T2D metaPRS was trained in a subset of 130,816 UK Biobank participants in the “White British” genetic ancestry cluster^15^. MetaPRS training comprised two key steps (**Figure 1**): (1) training of 44 individual component risk factor and diverse-ancestry T2D PRSs using LDpred2^26^, and (2) training the joint model combining the 44 T2D and related risk factor PRSs into a single meta-PRS using elasticnet penalized logistic regression^27^.

For step 1, we trained 44 PRSs for T2D using summary statistics from ten GWAS (or exome-wide association studies) for T2D across diverse ancestries and 34 GWAS for T2D risk factors (**Table S2**). To prevent overfitting, we selected contemporary GWAS that did not include UK Biobank participants^52^. The selected GWAS also did not include samples from any of the cohorts used for metaPRS evaluation in this study. Due to computational limitations of LDpred2, summary statistics were restricted to 1.6 million autosomal bi-allelic SNPs that were present in either the HapMap3 reference panel^53^ or in the two exome-wide association studies among the 44 GWASs (**Table S2**). When mapping GWAS summary statistics and HapMap3 variants to UK Biobank the UCSC Genome Browser^54^ liftOver tool was used to map positions from GRCh36 or GRCh38 to GRCh37 as needed. SNPs were further filtered on a per-GWAS basis following LDpred2 recommendations to remove variants with low power or divergent MAF between the GWAS and UK Biobank. LDpred2 was used to reweight GWAS summary statistics based on the linkage-disequilibrium of a subset of 11,074 UK Biobank participants enriched for T2D (1,202 cases) under multiple possible parameterisations of trait polygenicity and heritability (i.e. LDpred2 infinitesimal, grid-search, automatic, and lassosum models^26^). The remaining 120,464 UK Biobank participants (9,102 T2D cases) were then used to determine the optimal LDpred2 parameter choice for T2D prediction by assessing the AUC of logistic regression for combined prevalent and incident T2D case status (**Figure S1**, **Table S3**). Logistic regressions were fit adjusting for age and sex, and candidate PRSs were adjusted for 20 genetic PCs and standardized prior to model fitting.

Elasticnet penalized logistic regression^27^ was subsequently used in the 120,464 UK Biobank participants not used for LDpred2 parameter tuning to estimate the relative contributions of the 44 PRSs to T2D prediction and for deriving a single metaPRS (**Figure S2**, **Table S4**). The PC-adjusted 44 PRSs trained above were standardised and used as predictor variables along with age and sex in the regression. A range of elasticnet mixing parameters were tested (0, 0.1, 0.25, 0.5, 0.75, 0.9, and 1) with 10-fold cross-validation performed for each mixing parameter to tune the respective lambda penalty. The optimal regression fit was chosen as the combination of elasticnet mixing parameter and lambda penalty that had, across the 10-cross validation folds, the greatest mean AUC combined prevalent and incident T2D case status.

Per-SNP weights for the T2D metaPRS were subsequently derived via a weighted sum; where for each SNP *i*, the effect size was calculated as the sum of the per-SNP effect sizes γ derived from LDpred2 for each PRS *j* multiplied by the β coefficient estimated for the PRS in the optimal elasticnet regression:

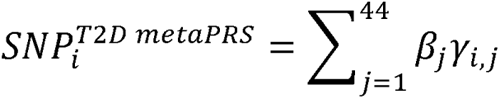

The T2D metaPRS comprised 1,349,896 SNPs, which we make available along with the respective weights through the PGS Catalog^9^ with accession PGS004923.

### Assessment of PRSs for prediction of T2D risk

For comparison with the metaPRS, genome-wide T2D PRS were obtained from PGS Catalog^9^ and separated into two groups: PRS whose training samples included UK Biobank participants of White European ancestries, and PRS whose training samples had no overlap with any of the cohorts analysed in this study (**Table S6**). PRS were calculated in each cohort as the weighted sum of their SNP weights multiplied by the dosages of the respective effect alleles. The Huerta-Chagoya et al. 2023 PRS was calculated as the weighted sum of their three component scores deposited as PGS003443, PGS003444, and PGS003445 following their formula of 0.531117×PGS003443 + 0.5690198×PGS003444 + 0.1465538×PGS003445 after standardising each component score to have mean 0 and standard deviation 1 in the target cohort^30^. Each PRS was subsequently adjusted for population structure by taking the residuals of a linear regression of the PRS on a cohort-specific number of genotype PCs (20 PCs for UK Biobank, INTERVAL, and All of Us).

Prediction of T2D case status at baseline assessment was assessed for each PRS separately using logistic regression adjusting for age at baseline and sex as covariates. In UK Biobank, assessment centre was also used as a covariate. PRS were standardized when fitting the regression so that reported odds ratios were per standard deviation increase and comparable across PRS. 95% confidence intervals for area under the receiver operating characteristic curves (AUC) were calculated using 2000 stratified bootstrap replicates^55^. Logistic regression was also used to assess incident T2D prediction in the Singapore Multi-Ethnic Cohort, as time to T2D onset (or T2D-free survival) was not available due to the heterogeneity of incident T2D ascertainment.

Prediction of incident T2D risk was assessed using Cox proportional hazards regression using time-in-study as the time scale and adjusting for age at baseline assessment and sex as covariates. Participants with T2D at baseline assessment were excluded. In UK Biobank, assessment centre was also used as a covariate. PRS were standardized when fitting the regression so that reported hazard ratios were per standard deviation increase and comparable across PRS. 95% confidence intervals for the Harrell’s C-index were calculated from the standard errors obtained using the infinitesimal jackknife method^56^.

### Comparison to established risk factors and risk scores

The metaPRS was compared to established T2D risk factors and 10-year T2D risk prediction scores (QDiabetes) in a subset of 190,293 UK Biobank participants from the metaPRS testing set that were free of T2D at baseline assessment and had quantified measurements for glucose and glycated haemoglobin (HbA1c).

We compared the metaPRS to the QDiabetes scores that are recommended for 10-year T2D risk prediction in the UK by the National Institute for Health and Care Excellence (NICE) guidelines in the UK^22^. Three QDiabetes scores are recommended depending on the availability of blood samples and fasting status: QDiabetes model A, which incorporates all risk factors that do not require taking a blood sample; QDiabetes model B, which additionally incorporates fasting glucose; and QDiabetes model C, which additionally incorporates HbA1c (but not fasting glucose). Risk factors used by all three QDiabetes models were participant age, sex, BMI, smoking status, Townsend deprivation index, family history of diabetes, antihypertensive medication usage, history of CVD, systematic corticosteroid medication usage, lipid lowering medication usage, history of gestational diabetes, history of polycystic ovary syndrome, history of learning difficulties, history of bipolar or schizophrenia disorders, and usage of 2^nd^ generation atypical antipsychotic medications. UK Biobank participants were non-fasting (median fasting time of 3 hours) so non-fasting glucose was used for QDiabetes model B. Details on each risk factor definition in UK Biobank are given in the **Supplementary Methods**.

Prediction of 10-year risk of incident T2D for each risk factor, risk score, and the metaPRS, were assessed using Cox proportional hazards regression using time-in-study as the time scale and adjusting for age, sex, and assessment centre. The metaPRS was adjusted for 20 genotype PCs prior to model fitting. Prior to model fitting BMI, glucose, and HbA1c were log transformed, and Townsend deprivation index was inverse rank normalized. All predictor variables were standardized when fitting Cox proportional hazard regressions. 95% confidence intervals for the Harrell’s C-index were calculated from the standard errors obtained using the infinitesimal jackknife method^56^.

Incremental improvement of the metaPRS over QDiabetes 2018 model C was assessed using multivariable Cox proportional hazards regression adjusting for age and sex. The change in C-index (ΔC-index) was calculated as the difference in C-index over a Cox proportional hazards regression fit for QDiabetes 2018 model C adjusting for age and sex. A bootstrap procedure with 1,000 bootstraps was used to estimate the standard error for the ΔC-index. Bootstrap resampling was performed using methods appropriate for right-censored data^57^. The 95% confidence interval and two-sided P-value were computed from the bootstrap standard error using the first order normal approximation method.

Incremental improvements in risk stratification when adding the metaPRS to QDiabetes risk scores (**Supplementary Methods**) were assessed at varying risk thresholds using categorical net reclassification improvement (NRI) analysis^58,59^. Categorical NRI analysis was used to assess relative to QDiabetes risk scores alone (1) the % of incident T2D cases correctly reclassified from low risk to high risk, and (2) the % of non-cases correctly reclassified from high risk to low risk. Bootstrap resampling of the categorical NRI analysis was performed using the nricens R package version 1.6, and 95% confidence intervals and P-values were subsequently calculated from the bootstrap standard error using the first order normal approximation method.

## Data Availability

Data from UK Biobank are available for health-related research subject to approval from the UK Biobank access committee. See https://www.ukbiobank.ac.uk/enable-your-research/apply-for-access for further details.
Data from the INTERVAL cohort can be requested by researchers for health-related research subject to approval from the INTERVAL Data Access Committee. Data will be shared through an institutional data sharing agreement. The INTERVAL Data Access Committee can be contacted via email at. Further information on the data access policy can be found at http://www.donorhealth-btru.nihr.ac.uk/project/bioresource.
Data from All of Us are available to researchers via the All of Us research hub subject to institutional data sharing agreement. For more information, see https://allofus.nih.gov/get-involved/opportunities-researchers.
Data from the Singapore Multi-Ethnic Cohort study can be requested by researchers for scientific purposes through an application process at the listed website (https://blog.nus.edu.sg/sphs/data-and-samples-request/). Data will be shared through an institutional data sharing agreement.

https://www.ukbiobank.ac.uk/enable-your-research/apply-for-access

https://allofus.nih.gov/get-involved/opportunities-researchers

https://blog.nus.edu.sg/sphs/data-and-samples-request/

http://www.donorhealth-btru.nihr.ac.uk/project/bioresource

## Data Availability

Data from UK Biobank are available for health-related research subject to approval from the UK Biobank access committee. See https://www.ukbiobank.ac.uk/enable-your-research/apply-for-access for further details.

Data from the INTERVAL cohort can be requested by researchers for health-related research subject to approval from the INTERVAL Data Access Committee. Data will be shared through an institutional data sharing agreement. The INTERVAL Data Access Committee can be contacted via email at. Further information on the data access policy can be found at http://www.donorhealth-btru.nihr.ac.uk/project/bioresource.

Data from All of Us are available to researchers via the All of Us research hub subject to institutional data sharing agreement. For more information, see https://allofus.nih.gov/get-involved/opportunities-researchers.

Data from the Singapore Multi-Ethnic Cohort study can be requested by researchers for scientific purposes through an application process at the listed website (https://blog.nus.edu.sg/sphs/data-and-samples-request/). Data will be shared through an institutional data sharing agreement.

## Code Availability

Code underlying this paper are available at https://github.com/sritchie73/T2D_metaPRS_paper. This repository and specific release for this paper are permanently archived by Zenodo at https://doi.org/10.5281/zenodo.13362823.

## Acknowledgements

The authors are grateful to UK Biobank, INTERVAL, All of Us, and the Singapore Multi-ethnic cohort for access to data to undertake this study.

Participants in the INTERVAL randomized controlled trial were recruited with the active collaboration of NHS Blood and Transplant (www.nhsbt.nhs.uk), which has supported fieldwork and other elements of the trial. DNA extraction and genotyping were co-funded by the National Institute for Health and Care Research (NIHR), the NIHR BioResource (http://bioresource.nihr.ac.uk) and the NIHR Cambridge Biomedical Research Centre (BRC) (no. BRC-1215-20014). The academic coordinating centre for INTERVAL was supported by core funding from the NIHR Blood and Transplant Research Unit in Donor Health and Genomics (no. NIHR BTRU-2014-10024), UK Medical Research Council (MRC) (no. MR/L003120/1), British Heart Foundation (nos SP/09/002, RG/13/13/30194 and RG/18/13/33946) and the NIHR Cambridge BRC (no. BRC-1215-20014). A complete list of the investigators and contributors to the INTERVAL trial is provided in ref. 17. The academic coordinating centre thanks blood donor centre staff and blood donors for participating in the INTERVAL trial.

The All Of Us Research Program is supported by the National Institutes of Health, Office of the Director: Regional Medical Centers: 1 OT2 OD026549; 1 OT2 OD026554; 1 OT2 OD026557; 1 OT2 OD026556; 1 OT2 OD026550; 1 OT2 OD 026552; 1 OT2 OD026553; 1 OT2 OD026548; 1 OT2 OD026551; 1 OT2 OD026555; IAA #: AOD 16037; Federally Qualified Health Centers: HHSN 263201600085U; Data and Research Center: 5 U2C OD023196; Biobank: 1 U24 OD023121; The Participant Center: U24 OD023176; Participant Technology Systems Center: 1 U24 OD023163; Communications and Engagement: 3 OT2 OD023205; 3 OT2 OD023206; and Community Partners: 1 OT2 OD025277; 3 OT2 OD025315; 1 OT2 OD025337; 1 OT2 OD025276. In addition, the All of Us Research Program would not be possible without the partnership of its participants.

The MEC study is supported by individual research and clinical scientist award schemes from the National Medical Research Council (NMRC) and the Biomedical Research Council (BMRC) of Singapore, and infrastructure funding from the Singapore Ministry of Health (Population Health Metrics and Analytics PHMA), National University of Singapore and National University Health System, Singapore. The MEC whole-genome sequencing data made use of data generated as part of the Singapore National Precision Medicine (NPM) program funded by the Industry Alignment Fund (Pre-Positioning) (IAF-PP: H17/01/a0/007).

This work was performed using resources provided by the Cambridge Service for Data Driven Discovery (CSD3) operated by the University of Cambridge Research Computing Service (www.csd3.cam.ac.uk), provided by Dell EMC and Intel using Tier-2 funding from the Engineering and Physical Sciences Research Council (capital grant EP/P020259/1), and DiRAC funding from the Science and Technology Facilities Council (www.dirac.ac.uk).

This work was supported by core funding from the British Heart Foundation (RG/18/13/33946: RG/F/23/110103), NIHR Cambridge Biomedical Research Centre (NIHR203312) [*], BHF Chair Award (CH/12/2/29428) and by Health Data Research UK, which is funded by the UK Medical Research Council, Engineering and Physical Sciences Research Council, Economic and Social Research Council, Department of Health and Social Care (England), Chief Scientist Office of the Scottish Government Health and Social Care Directorates, Health and Social Care Research and Development Division (Welsh Government), Public Health Agency (Northern Ireland), British Heart Foundation and the Wellcome Trust.

*The views expressed are those of the authors and not necessarily those of the NIHR or the Department of Health and Social Care.

L.P. was supported by a Rutherford Fund Fellowship from the Medical Research Council grant MR/S003746/1. X.J. was funded by British Heart Foundation (CH/12/2/29428) and Wellcome Trust (227566/Z/23/Z). Y.X. and M.I. are supported by the UK Economic and Social Research Council (ES/T013192/1). L. K. is funded by the NIHR BTRU in Donor Health and Behaviour (NIHR203337) and a BHF Chair award (CH/12/2/29428). S.A.L. was supported by a Canadian Institutes of Health Research postdoctoral fellowship (MFE-171279). F.S.C. acknowledges support from United States’ National Institutes of Health (NIH) grant ZIA-HG000024. J.C.D. acknowledges support from United States’ National Institutes of Health (NIH) grant ZIA-HG200417. J.D. holds a BHF Professorship and a NIHR Senior Investigator Award. E.D.A. holds a NIHR Senior Investigator Award. M.I. is supported by the Munz Chair of Cardiovascular Prediction and Prevention and the NIHR Cambridge Biomedical Research Centre (NIHR203312).

The funders had no role in study design, data collection and analysis, decision to publish, or preparation of the manuscript.

