## Supplementary Information for "Integrated clinical risk prediction of type 2 diabetes with a multifactorial polygenic risk score"

### Supplementary Figures

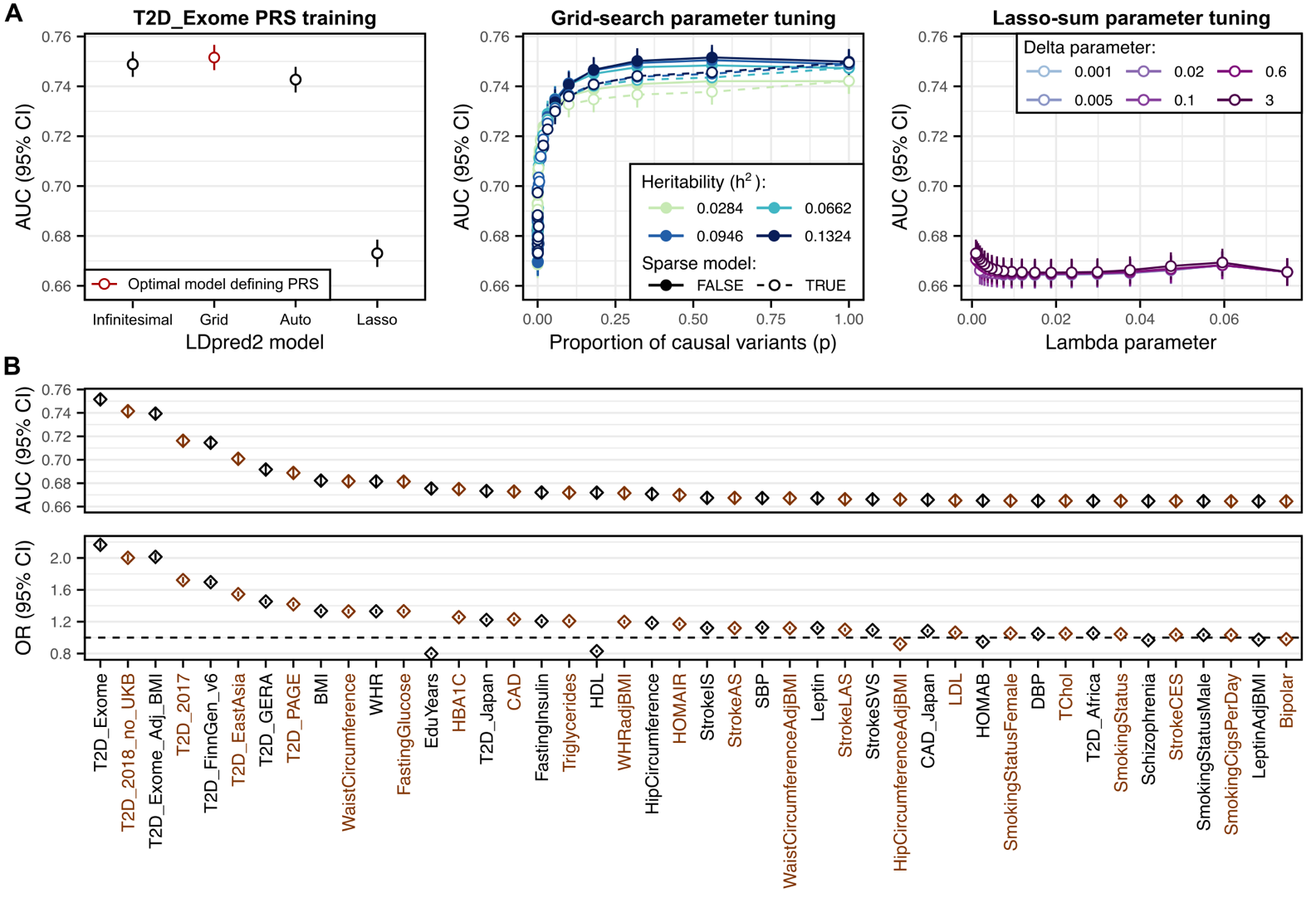

Figure S1: PRS training using LDpred2 for 44 PRSs underlying the T2D metaPRS

**A)** Selection of the optimal LDpred2 model and parameters when training the T2D_Exome PRS. Candidate PRSs were calculated in 120,464 UK Biobank participants (9,102 T2D cases) based on SNP weights estimated by LDpred2 in an independent set of 11,074 UK Biobank participants. AUC was assessed in the 120,464 participants after adjusting the candidate PRS for 20 genotype PCs and fitting an age- and sex- adjusted logistic regression for T2D. Four models of GWAS trait heritability and polygenicity were fit by LDpred2: infinitesimal, grid-search, automatic, and lassosum (**Methods**). The grid-search and lassosum models included multiple possible combinations of parameters. **B)** Area under the receiver-operating characteristic curve (AUC) and odds ratio (OR) for the optimal LDpred2 model and parameters selected when training each of the 44 PRSs underlying the T2D metaPRS. Diamonds show the AUC or odds ratios, and vertical bars show the 95% confidence intervals. Odds ratios for T2D are per standard deviation increase in the PC-adjusted PRS in age- and sex- adjusted logistic regression fit for each PRS separately. Details on the GWAS summary statistics underlying each PRS are provided in **Table S2**. Details on the LDpred2 model and parameter selection for each PRS are provided in **Table S3**.

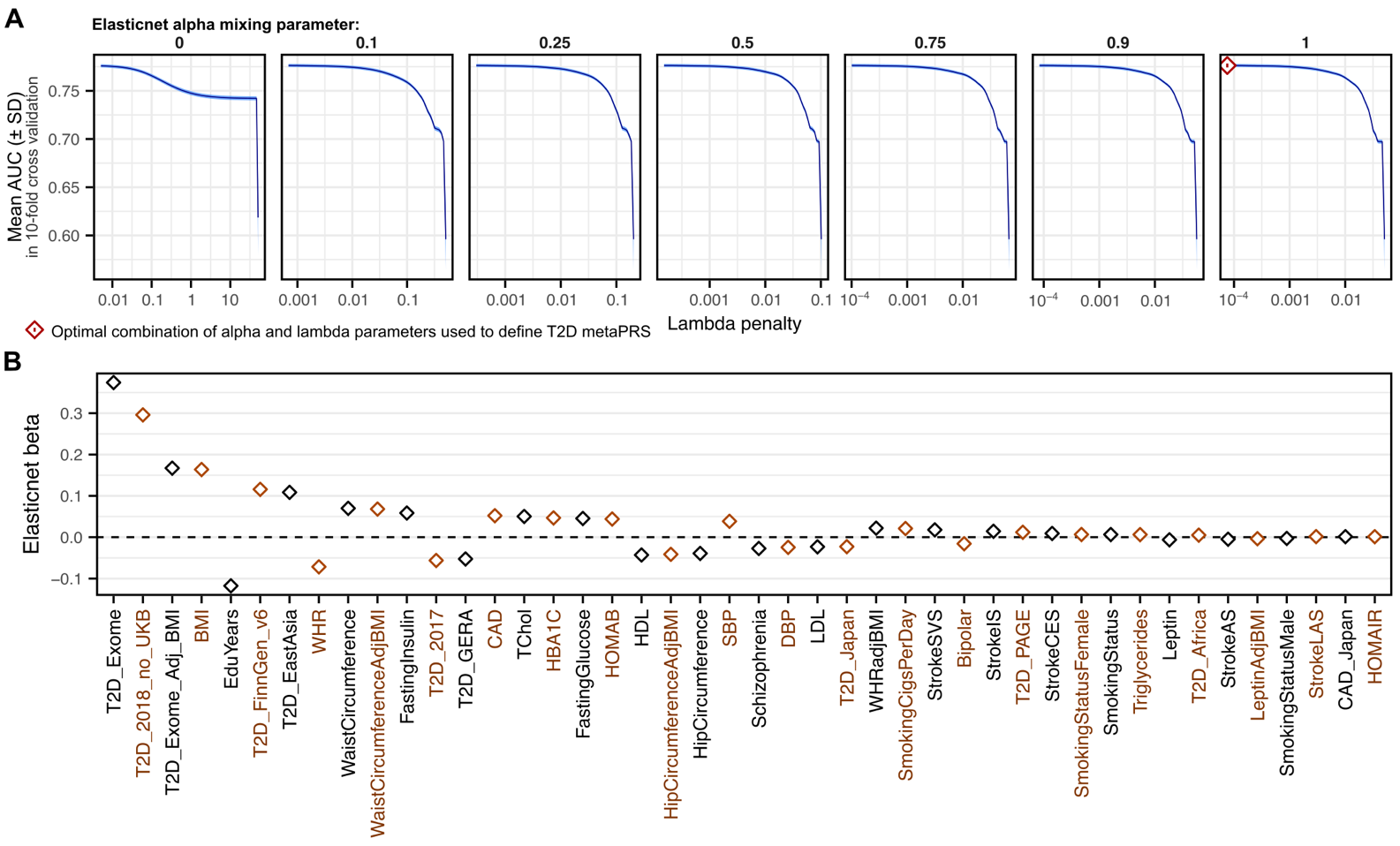

Figure S2: T2D metaPRS training

**A)** Elasticnet penalized logistic regression for T2D case status in 120,464 UK Biobank participants (9,102 T2D cases) with the 44 LDpred2-trained PRSs as predictor variables. Individual PRSs were adjusted for 20 genotype PCs prior to model fitting, and the elasticnet-penalized logistic regression were fit in 10-fold cross validation with genetic sex, baseline age, and assessment centre covariates. Each panel shows the mean AUC in 10-fold cross-validation (blue line) plus or minus the standard deviation (blue ribbon) for the given alpha mixing parameter value (panel header) and lambda penalty (x-axis). The red point highlights the combination of parameters that maximized the mean AUC (AUC: 0.776; SD: 0.00158) and were used to define the T2D metaPRS. Elasticnet penalized logistic regression were adjusted for genetic sex. **B)** Beta coefficients from the optimal elasticnet regression giving the relative contributions of each of the 44 component PRSs to the T2D metaPRS. Relative contributions of each PRS are detailed in **Table S4**.

### Supplementary Tables

Table S1: Characteristics of each cohort and genetic ancestry

In UK Biobank incident and prevalent T2D were analysed separately due to significant difference in phenotype severity: prevalent events were ascertained based on self-reported medical history and current medication usage, whereas incident events were ascertained from hospital records, thus only captured new cases that were severe enough to contribute to another disease event requiring hospitalisation. *Smoking status was available for only 73% of Singapore Multi-Ethnic Cohort participants. ^†^The metaPRS was trained on prevalent and incident cases combined, a breakdown of the cohort split into prevalent cases (and controls) and incident cases (and controls) is also shown.

Table S2: GWASs used to derive the T2D metaPRS

For each PRS underlying the T2D metaPRS, provides details on the GWAS trait, publication, sample size and case numbers, genetic ancestries, and source of summary statistics. See the Supplementary Reference for full details on each citation.

Table S3: PRS training using LDpred2 for 44 PRSs underlying the T2D metaPRS

Details the optimal LDpred2 model and parameters when training each PRS. SNPs: number of SNPs with non-zero weights in the PRS. LDpred2 model: optimal model of trait polygenicity and heritability fit by LDpred2 during PRS training. LDpred2 model parameters: respective model parameters (dependent on model type). AUC used for LDpred2 model and parameter selection was assessed in the 120,464 UK Biobank participants (9,102 T2D cases) after adjusting the candidate PRS for 20 genotype PCs and fitting an age- and sex- adjusted logistic regression for T2D. Odds ratios for T2D are per standard deviation increase in the PC-adjusted PRS. LDpred2 model parameters were estimated when reweighting the respective GWAS summary statistics (**Table S2**) using LDpred2 based on linkage-disequilibrium patterns in an independent set of 11,074 UK Biobank participants.

Table S4: Relative contributions of the 44 PRSs to the T2D metaPRS

Beta estimates from the optimal elasticnet regression in T2D metaPRS model training in 120,464 UK Biobank 1KG-EUR-like participants (9,102 T2D cases) giving the relative contributions of each of the 44 component PRSs to the T2D metaPRS. Beta estimates are per standard deviation increase in the respective PRS. Individual PRSs were adjusted for 20 genotype PCs prior to model fitting, and the elasticnet-penalized logistic regression were fit in 10-fold cross validation with genetic sex, baseline age, and assessment centre covariates. The optimal elasticnet regression whose beta coefficients are detailed here had a mean AUC of 0.776 with standard deviation of 0.00158 in 10-fold cross-validation.

Table S5: Comparison PRSs

Details on genome-wide T2D PRS obtained from the PGS Catalog for comparison to the metaPRS. The score development ancestries and underlying GWAS ancestries were obtained from the PGS Catalog entry for each T2D PRS. A column of yes/no values is used to flag whether the GWAS summary statistics underlying the PRS of score development of the PRS included UK Biobank samples. In all cases where this overlap occurred, the overlapping samples comprised only UK Biobank samples of European ancestries. There was no overlap between PRS training samples and evaluation cohort samples for any other cohort utilized in this paper. The Huerta-Chagoya *et al.* 2023 PRS was calculated as the weighted sum of their three component scores (PGS003443, PGS003444, and PGS003445) following their formula of 0.531117×PGS003443 + 0.5690198×PGS003444 + 0.1465538×PGS003445 after standardising each component score to have mean 0 and standard deviation 1 in the target cohort. See the Supplementary Reference for full details on each citation not appearing in the main text.

Table S6: Comparison of T2D PRSs in people of 1KG-EUR-like genetic ancestries across three cohorts

Data underlying Figure 2. Comparison of PRSs for predicting prevalent T2D status and/or incident T2D status in 1KG-EUR-like participants from the UK Biobank, INTERVAL, and All of Us cohorts. PRSs were adjusted for 20 genetic principal components in each cohort and genetic ancestry prior to model fitting. Odds ratios and hazard ratios are per standard deviation increase in the respective PC-adjusted PRS. Logistic and Cox proportional hazards regressions were adjusted for age, sex, and cohort specific covariates (e.g., assessment centre). Analyses of UK Biobank excluded participants used for metaPRS training, and PRSs derived from GWAS performed in UK Biobank samples of European genetic ancestries.

Table S7: Comparison of T2D PRSs in people of five diverse ancestry groups

Data underlying **Figure 3**. Comparison of PRSs for predicting prevalent T2D status and/or incident T2D in 1KG-AFR-like, 1KG-AMR-like, 1KG-EAS-like, 1KG-SAS-like, and ASN-like participants in UK Biobank, the All of Us research program, and Singapore Multi-Ethnic Cohort. PRSs were adjusted for 20 genetic principal components in each cohort and genetic ancestry prior to model fitting. Odds ratios and hazard ratios are per standard deviation increase in the respective PC-adjusted PRS. Logistic and Cox proportional hazards regressions were adjusted for age, sex, and cohort specific covariates (e.g., assessment centre). Logistic regression was used to assess incident T2D prediction in the Singapore Multi-Ethnic Cohort, as time to T2D onset (or T2D-free survival) was not available due to the heterogenous method of incident T2D ascertainment. Note the Shim *et al.* 2023 PRS (PGS003867) could not be tested in UK Biobank as it was derived from multi-ancestry GWAS performed in UK Biobank samples.

Table S8: Comparison of T2D metaPRS to established risk factors and risk scores

Data underlying **Figure 5**. Associations between individual risk factors and combined risk factor scores with 10-year risk of incident T2D in 190,293 1KG-EUR-like UK Biobank participants, selected for comparison purposes as the set of participants for whom the QDiabetes 2018 model C risk score could be computed (complete risk factor information, with height between 1.4 and 2.1 meters, weight ≤ 180 kg, and HbA1c between 15 and 48 mmol/mol). For models incorporating glucose a smaller number of participants was available for analysis. Additionally, QDiabetes 2018 model B required fasting glucose to be between 2 and 7 mmol/L. Note UK Biobank participants are non-fasting. Cox proportional hazards regressions were adjusted for age, sex, and UK Biobank assessment centre. Hazard ratios are reported for key predictor variables of interest for each model. Hazards ratios are per standard deviation increase for continuous predictors. The metaPRS was adjusted for 20 genetic principal components prior to model fitting. Where reported, the ΔC-index is relative to the respective QDiabetes risk score before adding the metaPRS. The 95% confidence intervals and P-values for the ΔC-index were computed in a bootstrap procedure with 1,000 bootstraps.

Table S9: Categorical net reclassification of 10-year risk with QDiabetes plus T2D metaPRS

Data underlying and extending **Figure 5C**. Details numbers stratified into low- and high- risk groups at varying risk thresholds when predicting risk with QDiabetes risk scores with or without the metaPRS, and numbers correctly reclassified and categorical net reclassification improvement (NRI) comparing risk scores with the metaPRS to those without the metaPRS. **A)** Stratification of future incident T2D cases into low- and high- risk groups, reclassification when comparing models with or without the metaPRS, and categorical NRI quantifying the % of cases correctly reclassified from low to high risk when adding the metaPRS to the QDiabetes model. 95% confidence intervals and P-values were calculated from the bootstrap standard error estimated via a bootstrap sampling procedure with 1000 bootstraps. **B)** As above, but for non-cases. Categorical NRI quantifies the % of non-cases correctly reclassified from high to low risk when adding the metaPRS to the QDiabetes model.

Table S10: Risk stratification and reclassification after initial screening with QDiabetes model A

Data underlying and extending **Figure 5D**. Details numbers stratified into low- and high- risk groups, and numbers correctly reclassified and categorical net reclassification improvement (NRI) comparing risk scores with the metaPRS to those without the metaPRS in two-stage risk screening: first with QDiabetes model A (with or without the metaPRS) to prioritize potential high risk individuals for fasting glucose or HbA1c blood tests (>5.6% risk following NICE and NHS health check guidelines), then second, risk-stratification at varying risk thresholds after predicting 10-year risk with QDiabetes models B or C (with or without the metaPRS) in those identified as potential high-risk by the initial screening step. Rows in blue show the numbers stratified by the initial screening step and case/non-case reclassification and categorical NRI comparing QDiabetes model A to QDiabetes model A plus the metaPRS. Subsequent non-shaded rows show the total numbers stratified after applying the second risk stratification step and compare the case/non-case reclassification and categorical NRI comparing the respective QDiabetes models with or without the metaPRS. *: 13.2% and 14.3% risk thresholds correspond to those recommended by the QDiabetes model development authors for risk stratification with models B and C respectively. **A)** Stratification of future incident T2D cases into low- and high- risk groups, reclassification when comparing models with or without the metaPRS, and categorical NRI quantifying the % of cases correctly reclassified from low to high risk when adding the metaPRS to the QDiabetes model. 95% confidence intervals and P-values were calculated from the bootstrap standard error estimated via a bootstrap sampling procedure with 1000 bootstraps. **B)** As above, but for non-cases. Categorical NRI quantifies the % of non-cases correctly reclassified from high to low risk when adding the metaPRS to the QDiabetes model.

### Supplementary Methods

#### Risk factor definitions in UK Biobank

Risk factors in UK Biobank were defined for consistency with and use for computation of QDiabetes risk scores^22^.

Age, sex, height, weight, body mass index, and Townsend deprivation index were extracted directly from fields #31, #21003, #50, #21002, #21001, and #189 respectively with no additional post-processing.

Smoking status was defined based on a combination of self-reported current smoking status (field #20116) and for current smokers, the self-reported number of cigarettes smoked per day (field #3456). Light smokers were those smoking <10 cigarettes per day. Moderate smokers were those smoking <20 cigarettes per day. Heavy smokers were those smoking ≥20 cigarettes per day. Smokers who answered “do not know” or “prefer not to answer” to the number of cigarettes smoked per day were grouped with moderate smokers.

Glucose (field #30740) and HbA1c (field #30750) concentrations above or below reportable assay limits (reported in fields #30746 and #30756 respectively) were set to the maximum and minimum values present in UK Biobank with a ±0.0001 offset. Note the median fasting time (field #74) was 3 hours and only 3% of participants were fasted for ≥8 hours at sample collection.

Family history of diabetes was ascertained based on self-reported information on family history in first-degree relatives (biological father, mother, or siblings; fields #20107, #20110, and #20111). Family history of diabetes was set to missing where it was not possible to determine presence or absence of diabetes in first-degree relatives based on the combination of “prefer not to answer” or “do not know” in these fields or where data were otherwise missing (e.g. there were no data on any of the touchscreen survey questions).

History of cardiovascular diseases (CVD) was determined from a combination of self-reported medical history from touchscreen questionnaires and verbal interview questions as well as retrospective hospital episode records. History of CVD included ischaemic heart disease, stroke, and transient ischaemic attacks. From the touchscreen survey (field #6150), this included self-reported heart attack or stroke. From the verbal interview with a nurse (fields #20002 and #20004), this included heart attack/myocardial infarction, cardiomyopathy, stroke, subarachnoid haemorrhage, brain haemorrhage, ischaemic stroke, transient ischaemic attack, coronary angioplasty (with or without stent), coronary artery bypass graft, or triple heart bypass. From linked hospital episode records this included coronary artery diseases (ICD-10 codes I21–I24 or ICD-9 codes 410–412), chronic ischaemic diseases (ICD-10 codes I25.1, I25.2, or I25.5–I25.9 or ICD-9 codes 414.0 or 414.8), any type of stroke (ICD-10 codes I60, I61, I63, or I64, or ICD-9 codes 430, 431, 434, or 436), or history of coronary surgery (ICD-10 code Z95.1, ICD-9 code V45.81, OPCS-4 codes K40-K46, K49, K50.1, or K75, or OPCS-3 codes 309.4 or 884). History of CVD was set to missing where it was not possible to determine presence or absence of history of CVD. This included people who did not satisfy any of the case definitions described above who had no data or reported “do not know” or “prefer not to answer” to any of the relevant self-report touchscreen survey or verbal interview questions.

History of polycystic ovary syndrome (PCOS) was determined in women based on a combination of self-reported medical history from verbal interview questions (field #20002) and retrospective hospital episode records (ICD-10 code E82.2 or ICD-9 code 256.4). Data were set to missing where it was not possible to determine presence or absence of PCOS. This included women who did not satisfy any of the case definitions described above who had no data or reported “do not know” or “prefer not to answer” to the relevant verbal interview questions.

History of gestational diabetes was determined in women based on a combination of self-reported medical history and retrospective hospital episode records using the Eastwood *et al.* algorithms^28^. Women with history of gestational diabetes were those who self-reported gestational diabetes in the touchscreen survey (field #4041) or in verbal interview (field #20002) with an age of diagnosis <50 years of age (field #20008) who did not concurrently self-report a history of type 1 or type 2 diabetes and also did not report taking any diabetes medications (fields #6177, #6153, #2986, and #20003; insulin products, metformin, sulfonylureas, meglitinides, glitazones, acarbose, glucobay, or glucotard), or had history of gestational diabetes in their hospital records (ICD-10 O24.2) or diabetes complications in pregnancy (ICD-10 024.3 or ICD-9 648.0, 648.8) with no record of any other type of diabetes (ICD-10 E10, E11, E13, E14, O24.0, O24.1 or ICD-9 250.00, 250.01, 250.09, 250.10, 250.11, 250.19, 250.29, or 250.99). Data were set to missing for women not classified as cases who had no data or reported “do not know” or “prefer not to answer” to the relevant touchscreen survey or verbal interview questions.

History of bipolar or schizophrenia disorders were determined from a combination of self-reported medical history from touchscreen questionnaires and verbal interview questions as well as retrospective hospital episode records. These included participants classified by Smith *et al.* 2013 as having type I or type II bipolar (data field #20122)^97^, self-reporting schizophrenia or mania/bipolar disorder/manic depression in the verbal interview (field #20002), or with history hospital episodes for bipolar disorder (ICD-10 code F31 or ICD-9 codes 296.80 or 296.89), schizophrenia (ICD-10 code F20 or ICD-9 code 295), or manic depressive psychosis (ICD-9 codes 296.0 or 296.4–296.7). Data were set to missing for women not classified as cases who had no data or reported “do not know” or “prefer not to answer” to the relevant touchscreen survey or verbal interview questions.

History of learning difficulties, a required risk factor for calculating QDiabetes scores^22^, was determined based on incidental records in historic hospital record linkage as no other information was collected by UK Biobank on this topic. Participants were classified as having possible history of learning difficulties if they had any record with ICD-10 codes F70–F79 or F81, or ICD-9 codes 315, or 317–319. An additional caveat here is that the hospital record linkage rarely extended back to childhood: baseline assessment occurred from 2006–2010 with mean age at assessment of 57 years, whilst the earliest retrospective hospital records available are from July 1993 for the 88% participants based in England, or April 1991 and December 1980 for the participants based in Wales (4% participants) and Scotland (8% participants) respectively.

Lipid-lowering medication usage was determined based on self-reported medication usage from touchscreen questionnaires (field #6177 for men, field #6153 for women) and in-person interview where a trained nurse reviewed and coded currently used or prescribed medications (field #20003). Participants were classified as using lipid lowering medications if they reported taking cholesterol lowering medication in the touchscreen survey or were recorded as taking any of the following medications: acipimox, atorvastatin, bezafibrate, bezafibrate product, ciprofibrate, colestipol, colestyramine, colestyramine product, colestyramine + aspartame 4g/sachet powder, exetimibe, fenofibrate, fluvastatin, gemfibrozil, gemfibrozil product, nicotinic acid product, pravastatin, rosuvastatin, or simvastatin. This list of lipid-lowering medications was determined by cross-referencing the UK Biobank medication list against the British National Formulary (BNF) chapter 2.12 lipid-regulating drugs (https://openprescribing.net/bnf/0212/) whose indications included lipid lowering for CVD prevention. Data were set to missing if the participant was not classified as case using the criteria above and either had no data recorded or answered “do not know” or “prefer not to answer” to the relevant touchscreen survey or verbal interview questions.

Hypertension medication usage was determined based on self-reported medication usage from touchscreen questionnaires (field #6177 for men, field #6153 for women) and in-person interview where a trained nurse reviewed and coded currently used or prescribed medications (field #20003). Participants were classified as using hypertension medications if they reported taking blood pressure medication in the touchscreen survey or were recorded as taking any of the following medications: bosentan, hydralazine, minoxidil, sildenafil, tadalafil, clonidine, clonidine hydrochloride 25 micrograms tablet, methyldopa, methyldopa + hydrochlorothiazide 250mg/15mg tablet, alpha methyldopa, moxonidine, guanethidine, doxazosin, indoramin, indoramin product, prazosin, terazosin, candesartan cilexetil, captopril, captopril + hydrochlorothiazide 25mg/12.5mg tablet, cilazapril, co-zidocapt 25mg/12.5mg tablet, enalapril, enalapril maleate + hydrochlorothiazide 20mg/12.5mg tablet, eprosartan, fosinopril, imidapril hydrochloride, irbesartan, irbesartan + hydrochlorothiazide 150mg/12.5mg tablet, lisinopril, lisinopril + hydrochlorothiazide 10mg/12.5mg tablet, losartan, losartan potassium + hydrochlorothiazide 50mg/12.5mg tablet, moexipril, olmesartan, amlodipine, perindopril, perindopril + indapamide, quinapril, ramipril, felodipine + ramipril, telmisartan, telmisartan + hydrochlorothiazide 40mg/12.5mg tablet, trandolapril, trandolapril + verapamil hydrochloride, verapamil, valsartan, or valsartan + hydrochlorothiazide 80mg/12.5mg tablet. This list of hypertension medications was determined by cross-referencing the UK Biobank medication list against the BNF chapter 2.5 Hypertension and heart failure drugs (https://openprescribing.net/bnf/0205/) whose indications included hypertension. Data were set to missing if the participant was not classified as case using the criteria above and either had no data recorded or answered “do not know” or “prefer not to answer” to the relevant touchscreen survey or verbal interview questions.

Systematic corticosteroid medication usage was determined based on self-reported medication usage from the in-person interview where a trained nurse reviewed and coded currently used or prescribed medications (field #20003). Participants were classified as using systematic corticosteroids if they were recorded as taking any of the following medications: betamethasone, deflazacort, dexamethasone, hydrocortisone, methylprednisolone, prednisolone, prednisolone product, prednisone, or triamcinolone. This list of systematic corticosteroids medications was determined by cross-referencing the UK Biobank medication list against the BNF chapter 6.3.2 Glucocorticoid therapy drugs (https://openprescribing.net/bnf/060302/) including only drugs that were taken orally or via injection as well as the list used by Hippisley-Cox and Coupland in their development of the QDiabetes risk scores^22^. Data were set to missing if the participant was not classified as case using the criteria above and either had no data recorded or answered “do not know” or “prefer not to answer” to the relevant touchscreen survey or verbal interview questions.

Second generation atypical antipsychotic medication usage was determined based on self-reported medication usage from the in-person interview where a trained nurse reviewed and coded currently used or prescribed medications (field #20003). Participants were classified as using 2^nd^ generation atypical antipsychotics if they were recorded as taking any of the following medications: amisulpride, aripiprazole, clozapine, olanzapine, quetiapine, risperidone, sertindole, or zotepine. This corresponds to the list used by Hippisley-Cox and Coupland in their development of the QDiabetes risk scores^22^. Data were set to missing if the participant was not classified as case using the criteria above and either had no data recorded or answered “do not know” or “prefer not to answer” to the relevant touchscreen survey or verbal interview questions.

#### QDiabetes risk scores in UK Biobank

QDiabetes risk scores were calculated using the QDiabetes R package in 1KG-EUR-like UK Biobank participants not used for metaPRS training who had complete information and met requisite range restrictions on the risk factors composing each QDiabetes model^22^. In addition to non-missing data, the QDiabetes risk scores required participants to be ≥1.4 meters and ≤ 2.1 meters in height, and ≥40 kg and ≤180 kg in weight. Further participants within these ranges whose BMI were either <20 or >40 were winsorized to 20 or 40 respectively by the QDiabetes algorithm. For models incorporating either fasting glucose (model B) or HbA1c (model C) the QDiabetes risk scores required participants to have fasting glucose ≥ 2 and <7 mmol/L and HbA1c ≥ 15 and < 48 mmol/mol. Non-fasting glucose was used as a proxy when calculating 10-year risk using QDiabetes model B as fasting glucose was not available. Note the median fasting time (field #74) was 3 hours and only 3% of participants were fasted for ≥8 hours at sample collection, so it is expected that model B will systematically overestimate risk in UK Biobank, with varying degree of overestimation depending on the fasting time (and enrichment for missing data in people who have recently eaten).

#### Absolute risk prediction with QDiabetes risk scores plus T2D metaPRS

The T2D metaPRS was added to the each QDiabetes risk score (models A, B, and C) using the method described by Hageman *et al.* 2023 for flexible addition of risk modifiers to established risk scores^98^. The following formula was used:

$$1-\left( 1-QDiabetes absrisk \right)^exp\left( {logHR}_{metaPRS}\times{Zscore}_{metaPRS} \right)$$

The formula was applied in males and females separately as the QDiabetes risk scores have differing baseline hazards and risk factor weightings in males and females^22^. The Z-score represents the per-participant level of the T2D metaPRS adjusted for 20 genotype PCs then standardized in males and females separately. The log hazard ratio (logHR) was calculated for this Z-scored metaPRS using Cox proportional hazards regression fit in on the males and females separately adjusting for the linear predictor of the respective QDiabetes risk score as an offset term.
